# Single-port laparoscopic surgery for sigmoid colon cancer: a novel surgical strategy based on “peritoneal dome”

**DOI:** 10.1101/2024.12.16.24319085

**Authors:** Lv Qijun, Tang Jin, Ye Pengcheng, Shen Jun, Wang Qinyuan, Chen Jingjing, Huang Hongshen, Gan Hailin, Li Junyi, Lin Ruijie, Liu Yuhang, Wei Shoujiang

## Abstract

**Aims:** This study aimed to introduce a novel ‘peritoneal dome’ strategy for single-port laparoscopic surgery in patients with sigmoid colon cancer and to evaluate the feasibility and safety of this approach.

**Methods:** A total of 12 patients at the affiliated hospital of North Sichuan Medical College from January 2022 to August 2024 were enrolled. All procedures were performed by the same surgical team. By collecting and analyzing the demographic information, clinical characteristics, surgical outcomes and postoperative recovery indicators of this group of patients, the short-term efficacy and safety of this surgical strategy were preliminarily studied.

**Results:** The median age of patients was 72 years (range, 58-81), with a median body mass index (BMI) of 25.8 kg/m^2^ (range, 22–31.2). Most patients had underlying respiratory conditions, particularly chronic obstructive pulmonary disease (COPD) (80%). All patients successfully completed the single-port laparoscopic surgery without conversion to laparotomy. The median of the total operating time was 98.80 minutes. The mean arterial carbon dioxide pressure (PaCO2) during the extraperitoneal phase was significantly lower (32.5 mmHg) compared to the intraperitoneal phase (47.1 mmHg). The median estimated blood loss was 19.20 ml, with no transfusions required and no significant operative complications reported. Pathological analysis revealed that 83.3% of patients had advanced disease (stage III), and the average number of harvested lymph nodes was 18.92. Postoperatively, patients experienced a mean time to ambulation of 1.25 days and a mean hospital stay of 6.17 days. Complications occurred in 33.3% of patients, primarily classified as grade I and II according to the Clavien-Dindo classification. Notably, there were no cases of anastomotic leakage or severe complications.

**Conclusion:** The single-port laparoscopic surgery based on a “peritoneal dome” supported strategy for sigmoid colon cancer demonstrates effective oncological management with minimal complications and a favorable recovery profile. The results suggest that this technique is a viable option for patients with advanced sigmoid colon cancer, particularly in those with significant comorbidities.

## Introduction

As a remarkable global health problem, new cases of Colorectal cancer (CRC) was nearly 1.9 millions and reach the second leading cause of cancer-related deaths worldwide according to the Global Cancer Statistics 2022 [1]. Over the past two decades, the principles of minimally invasive surgery, particularly laparoscopic techniques, have become integral to the management of gastrointestinal tumors[2]. In recent years, numerous prospective randomized controlled trials have substantiated the safety and efficacy of laparoscopic surgery for colorectal cancer[3]. As the paradigm of minimally invasive surgery continues to evolve, single-incision laparoscopic surgery has emerged as a viable option for selected patients with gastrointestinal tumors. However, the technical complexity associated with single-incision laparoscopic procedures results in extended surgical durations[4]. This is particularly concerning in elderly patients[5,6], as prolonged exposure to CO2 pneumoperitoneum may precipitate hypercapnic acidosis[7], thereby complicating perioperative recovery and impeding the broader adoption of single-incision laparoscopic techniques.

In advanced colorectal cancer, the metastasis rate of lymph nodes along the root of the inferior mesenteric artery (No. 253 LN) ranges from 0.3% to 14.2%[8], while the metastasis rate for para-aortic lymph nodes (PALNs) is approximately 2%[9]. Notably, the pathological positivity rate for PALNs in patients with preoperative imaging lymphadenopathy is 37.8%[10]. Therefore, for select cases of sigmoid colon and rectal cancer, clearance of No. 253 lymph nodes is recommended[11]. Although routine clearance of PALNs is not advised, preoperative imaging has demonstrated a high metastasis rate in PALNs with a long axis exceeding 0.7 cm, indicating that these patients should undergo PALN clearance[10].However, the technical challenges of clearing PALNs during laparoscopic surgery are significant, prompting surgeons to explore various surgical strategies. Vasilev and McGonigle first described laparoscopic extraperitoneal para-aortic lymph node dissection in 1996[12], demonstrating its safety and feasibility. Theoretically, combining laparoscopic techniques with an extraperitoneal approach may reduce complications such as intestinal injury and adhesions, potentially shortening hospital stays[13]. Nonetheless, conventional laparoscopic methods face considerable difficulties in PALN clearance due to intestinal interference, especially under single-incision laparoscopic conditions where instrument overlap complicates exposure of the para-aortic region near the renal vein[14]. In gynecologic oncology, surgeons have employed a “tent-pitching” dissection technique to minimize small intestinal interference[15]. While this method eliminates the need for assistance, it poses challenges due to the opposing camera view, resulting in a steep learning curve and making it impractical in single-incision laparoscopic settings[15]. Efetov and colleagues also reported a case of right colon resection by retroperitoneal approach[16]. Under single-incision laparoscopic conditions, the traction from a wound retractor allows for seamless internal-external conversion, enabling us to suture the temporarily opened posterior peritoneum to the anterior abdominal wall and facilitating access to the retroperitoneum.Additionally, low-pressure CO2 injection through the single-incision laparoscopic retractor elevates the retroperitoneal space, creating a “peritoneal dome” without the need for assistant traction, thus achieving excellent retroperitoneal exposure. Based on this, we proposes a novel surgical strategy based on “peritoneal dome” to create an effective operation space retroperitoneally for lymph node dissection and vascular dissection. Herein, this surgical strategy was used for 12 patients with sigmoid colon cancer, the feasibility and safety of this surgical strategy also evaluated.

## Materials and methods

### Patient population

This retrospective study enrolled 12 patients with advanced sigmoid colon cancer. And patients underwent a single-port laparoscopic local extraperitoneal mixed approach in the affiliated hospital of North Sichuan Medical College from January 2022 to August 2024. All surgical procedures are performed by the same surgical team.

### Observed indicators

The characteristics of the patients included age, body mass index (BMI), tumor size, tumor stage, tumor grade, blood loss, extraperitoneal operation time, intraperitoneal operation time, mean arterial carbon dioxide pressure, intraoperative and postoperative complications, and recovery time of intestinal function, pathology, lymph node count, lymph node status, length of hospital stay and other indicators.

### Surgical approach

After anesthetized, the patient was placed in the lithotomy position. This operation needs two surgeons to complete, after the routine disinfection cloth, the surgeon standing on the right side of the patient, while the assistant is standing on the right side of the patient. A 3 cm longitudinal incision was made in the midline of the abdomen under the umbilical cord, and the layers of the abdominal wall were cut and entered the abdominal cavity. A single-hole laparoscopic retractor was used to retract the incision, and the CO2 pneumoperitoneum was established, keeping the intra-abdominal pressure less than 14 mmHg. A 30-degree laparoscope was introduced, and the peritoneal cavity was examined and the primary lesion was explored. When the exploration is complete, the patient takes the lower-foot-high lithotomy position and cuts the right peritoneum about 5 cm at the level of the Cape sacrum as a traditional laparoscopy. The peritoneum can be raised about 2 cm high by proper dissection. Closing the pneumoperitoneum, taking out the single-port laparoscopic retractor, suturing and fixing the opened lateral peritoneum to the peritoneum layer of the anterior abdominal wall with 4 needles, forming a “peritoneal dome” in the abdominal cavity, and then placing the single-port laparoscopic retractor again, and re-injection of CO2 gas, adjust CO2 pressure to 5 mmhg, can enter the retroperitoneal space. The root of the Inferior mesenteric artery was isolated and ligated along the loose laparoscopy tissue, and No.253LN were removed if necessary, the paraaortic lymph nodes can be removed to the level of renal artery, while the paraaortic lymph nodes can be removed to the posterorectal space to the level of sphincter groove Both sides may reach the basin wall. If lateral lymphadenectomy is required, the area of lateral lymphadenectomy may be entered along the internal iliac artery. After the retroperitoneal dissociation, the pneumoperitoneum can be discharged and the single-port laparoscopic retractor can be taken out, the 4 needles between the anterior and posterior peritoneum which were sutured before can be removed, and the CO2 pneumoperitoneum can be established, the peritoneum was excised and the distal rectum was excised in a relatively short time. The proximal colon can be pulled out of the body and placed on the stapler base for end-to-end colorectal anastomosis, as is traditionally laparoscopy.

### Statistical analysis

Statistical analysis using SPSS 22.0. Descriptive analysis was performed and demographics, clinical, pathological, and surgical variables were summarized. BMI is calculated by dividing your weight in kilograms by your height in meters squared. Median and interquartile ranges were calculated for continuous data, as well as percentages for categorical data. Quantitative data are presented as mean ± S.D. for normally distributed data.

## Results

This retrospective study enrolled a total number of 12 patients.All patients were diagnosed as sigmoid adenocarcinoma with a median distance of 14 cm (12-19 cm) from the anus by colonoscopy and pathological examination. All patients had no obvious gastrointestinal obstruction and acute bleeding. As shown by **Table 1**, the median age of the patients was 72 years (range, 58-81 years); the median BMI was 25.8 kg/m2 (range, 22–31.2 kg/m2). Four patients had received 2 cycles of neoadjuvant chemotherapy ( mFOLFOX6) before surgery. In addition, most of the cases in this study had underlying heart and lung disease, especially COPD, which occurred in 80% of patients.The clinical data of this group of patients showed that the average size of the tumor was 3.85cm and the clinical staging of stage I, II, and III patients was accounted for 8.3%,25%,66.7%, respectively. In particular, preoperative imaging results also suggested that the enlargement of No.253 lymph nodes were founded in all 12 patients.

**Table 1.** Clinical characteristics baseline of patients.

|  | Mean (range or %) |
| --- | --- |
| Age (year) | 72 ( 65-77) |
| Gender |  |
| Male | 7(58.3) |
| Female | 5(41.7) |
| BMI (kg/m<sup>2</sup>) | 27(20-32) |
| ASA score |  |
| II | 4 (33.3) |
| III | 6 (50) |
| IV | 2 (16.7) |
| Underlying medical conditions |  |
| COPD | 10(83.3) |
| Hypertension | 8 (66.7) |
| Diabetes | 4(33.3) |
| Clinical TNM stage |  |
| I | 1 (8.3) |
| II | 3 (25) |
| III | 8 (66.7) |
| Enlarged lymph nodes of No.253 | 12(100) |

In terms of surgical outcome (**Table 2**), all 12 patients successfully completed the single-port laparoscopy as planned, and no case was converted to laparotomy. The median operative time was 35 minutes (21-55 minutes) for extraperitoneal operation and 48 minutes (37-57 minutes) for intraperitoneal operation. Our results showed that retroperitoneal surgery with low CO2 pressure significantly reduced blood CO2 levels compared with conventional laparoscopy.Specifically, the mean value of PaCO_2_ was 32.5 mmHg and 47.1 mmHg during the period of extraperitoneal and intraperitoneal procedures, respectively. And the median blood loss for the entire procedure was 19.20 ml (10-35 ml), no blood transfusion during the operation was needed in all patients, and also no operative complications was occurred. In the present study,the results of the surgical evaluation indicated that the mean incision length was 3.83 cm and the colonic fascia propria remained intact across all cases. Other surgical outcomes were also assessed (**Table 2**), the proximal resection margin exhibited a mean length of 9.24 cm, while the distal resection margin measured 4.96 cm. Additionally, an average of 18.92 lymph nodes were harvested during the procedure. For the No. 235 group lymph nodes, a median of 3 lymph nodes was successfully harvested. Furthermore, PALNs dissection was conducted in three patients, yielding an average of 3 lymph nodes per patient, with a range of 0 to 6 lymph nodes collected. These findings suggest effective lymphatic management and underscore the adequacy of the surgical technique employed in ensuring proper oncological clearance.

**Table 2.** Surgical and pathological outcomes.

|  | N (%) or mean (SD) |
| --- | --- |
| Total operating time (min) | 98.80 (81-127) |
| Time of retroperitoneal operation (min) | 35 (21-55) |
| Time of intra-abdominal operation (min) | 48 (37-57) |
| Estimated blood loss (ml) | 19.2 (7.64) |
| Additional trocar | 1 (8.3) |
| Retroperitoneum rupture | 1 (8.3) |
| Arterial tension of carbon dioxide (PaCO<sub>2</sub>) |  |
| During retroperitoneal operation (mmHg) | 32.5(7.5) |
| During intra-abdominal operation (mmHg) | 47.1(9.3) |
| Incision length (cm) | 3.83 (0.83) |
| Tumor size (cm) | 3.85 (0.99) |
| Proximal resection margin (cm) | 9.24 (1.97) |
| Distal resection margin (cm) | 4.96 (0.75) |
| No. of harvested lymph nodes | 18.92 (2.68) |
| Differentiation |  |
| Well | 5 (41.7) |
| Moderate | 4(33.3) |
| Poor | 3 (25) |
| Depth of invasion |  |
| T1 | 3 (25) |
| T2 | 3 (25) |
| T3 | 5 (41.7) |
| T4 | 1 (8.3) |
| Lymph node metastasis |  |
| N0 | 2 (16.7) |
| N1 | 6 (50) |
| N2 | 4 (33.3) |
| Positive of NO.253 lymph nodes | 4(33.3) |
| Pathologic TNM stage |  |
| I | 1 (8.3) |
| II | 1 (8.3) |
| III | 10 (83.3) |

The pathological assessment of patients revealed varied differentiation levels within the these tumor samples. Specifically, 5 tumors (41.7%) were categorized as well-differentiated, 4 tumors (33.3%) as moderately differentiated, and 3 tumors (25%) as poorly differentiated. Regarding the depth of invasion, tumors of 3(25%) patients were classified as T1, 3 tumors (25%) as T2, 5 tumors (41.7%) as T3, and 1 tumor (8.3%) as T4. Lymph node involvement was noted, with 2 patients (16.7%) classified as N0, 6 patients (50%) as N1, and 4 patients (33.3%) as N2. Notably, a total of 4 out of 12 examined No.253 lymph nodes (33.3%) were positive among those patients.In terms of the pathologic TNM staging, there was a predominance of advanced disease, with 10 patients (83.3%) classified as stage III, while 1 patient (8.3%) was categorized as stage I and another as stage II. These findings indicate a significant burden of disease and suggest a need for careful postoperative management and monitoring in this patient population.

As shown in the **Table 3**, the postoperative recovery metrics revealed a mean time to ambulation of 1.25 days (SD 0.34). Patients experienced a mean time to first flatus of 1.63 days (SD 0.38) and a mean time to first defecation of 2.83 days (SD 1.25). The transition to a liquid diet occurred at a mean of 1.6 days (SD 0.57) postoperatively.For Pain assessment, which was measured by using the Visual Analog Scale (VAS), indicated scores of 2.58 (SD 0.79) on postoperative day 1 (POD 1), 2.42 (SD 0.67) on POD 2, 2.08 (SD 0.79) on POD 3, and a notable decrease to 0.83 (SD 0.72) by POD 5. And the overall postoperative hospital stay averaged 6.17 days (SD 1.38). These results suggest that these patients experience a relatively rapid recovery trajectory with manageable pain levels postoperatively.

**Table 3.** Morbidity, mortality, and recovery courses.

|  | N (%) or mean (SD) |
| --- | --- |
| Total perioperative complication | 4 (33.3) |
| Wound complication | 1 (8.3) |
| Anastomotic leakage | 0 (0.0) |
| Anastomotic bleeding | 0 (0.0) |
| Lymphorrhagia | 1 (8.3) |
| Intra-abdominal bleeding | 0 (0.0) |
| Intra-abdominal infection | 0 (0.0) |
| Bowel obstruction | 0 (0.0) |
| Cardiocerebral events | 0 (0.0) |
| Respiratory infection | 2 (17.6) |
| Clavien-Dindo classification |  |
| I | 1 (8.3) |
| II | 3 (25) |
| III | 0 (0.0) |
| Mortality | 0 (0.0) |
| Time to ambulation (day) | 1.25(0.34) |
| Time to first flatus (day) | 1.63 (0.38) |
| Time to first defecation (day) | 2.83 (1.25) |
| Time to liquid diet (day) | 1.6(0.57) |
| VAS |  |
| POD 1 | 2.58(0.79) |
| POD 2 | 2.42 (0.67) |
| POD 3 | 2.08 (0.79) |
| POD 5 | 0.83 (0.72) |
| Postoperative hospital stay (day) | 6.17 (1.38) |

In this study, a total of four patients with perioperative complications were observed (**Table 3**), corresponding to a complication rate of 33.3%. Specifically, wound complications occurred in 1 patient (8.3%), while lymphorrhagia was noted in another patient (8.3%). Notably, there were no cases of anastomotic leakage, anastomotic bleeding, intra-abdominal bleeding, intra-abdominal infection, bowel obstruction, or cardiocerebral events. Moreover, respiratory infections were identified in 2 patients (17.6%). According to the Clavien-Dindo classification, complications were categorized as follows: grade I complications were observed in 1 patient (8.3%), while grade II complications were noted in 3 patients (25%). No patients experienced grade III complications. These results indicate that while some complications were present, they were relatively few and manageable, suggesting an overall favorable postoperative outcome in our patient cohort.

With advanced disease burden, 10 patients were administered adjuvant chemotherapy following surgery, which commenced on a mean postoperative day of 17 days (range: 14-22 days). Comprehensive follow-up was conducted for all patients, yielding a median follow-up duration of 17 months, with a range of 1 to 30 months. The follow-up protocol included assessments at 1 month, 3 months, and 6 months post-surgery, followed by biannual evaluations.Importantly, all patients showed no signs of recurrence at the most recent follow-up, but long-term oncological outcomes require continued follow-up to be obtained.

## Discussion

Laparoscopy has emerged as the standard surgical approach for colorectal cancer, and advancements in surgical techniques have led to the adoption of single-port laparoscopy for select patients with gastrointestinal tumors[17]. Despite the progress made in some large medical centers, the widespread implementation of laparoscopy has been hindered by various technical challenges. One of the primary obstacles of single-port laparoscopy is the interference between the main surgical instrument and the assistant’s tools[18].To address this inherent limitation, it is crucial to rethink traditional approaches and strategies to minimize the assistant’s exposure requirements. Some intracavitary self-traction instruments and traction techniques have been developed to alleviate the reliance on assistants[19]; however, these solutions often exhibit limited flexibility and necessitate frequent adjustments in traction direction and positioning, thereby prolonging operative time and complexity.The introduction of single-port laparoscopic retractors offers a promising alternative. By facilitating the opening and suturing of the retroperitoneum to the anterior abdominal wall, these retractors can create a “peritoneal dome” that effectively isolates the small intestine and colon from the retroperitoneal space[16]. This innovative approach significantly reduces the need for assistant-operated devices, thereby mitigating some of the technical difficulties associated with single-port laparoscopy.

Furthermore, single-port laparoscopy presents significant technical challenges that often lead to increased operative time, particularly in elderly patients who are more susceptible to hypercapnia due to prolonged CO2 pneumoperitoneum. This condition can complicate perioperative recovery and poses a barrier to the widespread adoption of single-port laparoscopic techniques.To mitigate hypercapnia associated with pneumoperitoneum, two strategies can be employed: expediting the surgical procedure and either reducing pneumoperitoneum pressure or avoiding it altogether[20]. However, minimally invasive surgical techniques, such as pneumoperitoneum-free surgery and conventional multi-ports laparoscopic procedures conducted under low pneumoperitoneum pressures, encounter significant limitations primarily stemming from the challenge of maintaining adequate surgical field exposure[21]. The reduced maneuverability and operating arm limitations inherent in single-port laparoscopic surgery under low pneumoperitoneum conditions can significantly increase the surgical difficulty. Therefore, minimally invasive surgical modification methods based on the above two conventional strategies cannot address the problem of hypercapnia in high-risk patients.In this study, we implemented a single-port laparoscopic extraperitoneal hybrid approach, allowing the colon cancer resection to be performed retroperitoneally. This method enabled us to maintain pneumoperitoneum pressure at significantly lower levels (e.g., 4-5 mmHg), resulting in markedly reduced carbon dioxide levels in the patients’ blood compared to traditional laparoscopic techniques. Our findings demonstrated that the partial pressure of CO2 in arterial blood was only 32.5 mmHg during extraperitoneal procedures, in contrast to 47.1 mmHg during intraperitoneal procedures. These results suggest that this hybrid extraperitoneal approach is an effective method for reducing hypercapnia during laparoscopic surgery.

In cases of advanced colorectal cancer, regional lymph node dissection is essential when preoperative imaging indicates the presence of para-aortic lymph node or lateral lymphadenopathy[22]. However, traditional laparoscopic dissection of PALNs poses significant challenges due to interference from the small intestine, particularly in the context of single-port laparoscopy[23]. In this study, we employed a hybrid surgical approach that involved the introduction of a low-pressure CO2 insufflation into the retroperitoneal space. This technique facilitated a novel surgical strategy based on “peritoneal dome”, effectively mitigating interference from the small intestine. The insufflation of CO2 also allowed for the fat and lymphatic tissue around the aorta more flexible,which makes it easy to dissect the lymph nodes below the level of the renal vein.Furthermore, accurately identifying the appropriate incision point is crucial in cases necessitating lateral lymph node dissection[24]. The novel surgical approach presented in this study facilitates lymph node dissection by allowing access from the bifurcation of the iliac artery and along the branches of the internal iliac artery, thereby enhancing the overall efficiency and effectiveness of the procedure.

After conventional sigmoidectomy, adhesions often form between the exposed retroperitoneal tissue and the bowel due to excessive posterior peritoneal defects[25]. These adhesions can lead to complete or partial intestinal obstruction in a significant number of cases. Resuturing the opened peritoneal tissue postoperatively may reduce the risk of adhesion[26]; however, conventional laparoscopic techniques often necessitate the removal of more peritoneal tissue to ensure adequate surgical field exposure, which complicates the possibility of suture reconstruction.In contrast, the approach utilized in this study allows for the dissection of the peritoneum and mesentery while ensuring that the retroperitoneal tissue remains sufficiently intact. This not only facilitates the radical resection of the tumor but also provides a robust foundation for the reconstructing the peritoneal tissue and the coverage of the surgical wound.

Nevertheless, the indications for the single-port laparoscopic local extraperitoneal hybrid approach remain to be thoroughly defined. Similar to previous report, one notable advantage of this method is its ability to effectively manage the flat tissue of the abdominal aorta while adequately minimizing interference from intra-abdominal organs[16]. This suggests that a similar surgical paradigm could potentially be employed for other retroperitoneal lesions. Comparable research in gynecological oncology has demonstrated the feasibility of utilizing handheld devices to elevate the retroperitoneum under conditions of porous laparoscopy, allowing for complete dissection of retroperitoneal lymph nodes up to the level of the renal veins[15,23]. In previous study[23], the formation of the peritoneal “tent-pitching” relies on the continuous traction of the peritoneum by an assistant during the whole procedure.Differently, the methodology adopted in this study fundamentally leverages the properties of CO2 to elevate the intact peritoneum, thereby enhancing reliability compared to the techniques employed in previous studies. The supportive function of the gas simplifies tissue separation, making this modification analogous to the transition from transabdominal preperitoneal (TAPP) to totally extraperitoneal (TEP) approaches in the context of abdominal hernia repairs[27].

Based on our limited practical experience, several critical considerations must be addressed when utilizing the novel surgical strategy based on “peritoneal dome” in single-port laparoscopic surgery. After establishing the retroperitoneal access, it is imperative to accurately identify the appropriate anatomical plane to successfully enter the Toldt’s space, particularly posterior to the inferior mesenteric artery.Moreover, due to the inherent limitations of the retroperitoneal perspective, there is often a lack of direct visualization to ensure the tumor site. Therefore, after identifying the tumor lesion during intra-abdominal exploration, it is essential to utilize markers such as methylene blue or nano-carbon to delineate the predetermined resection points. This marking facilitates the guidance of the extent of retroperitoneal dissection.Another important aspect of this technique is ensuring the integrity of the retroperitoneum during the dissection process. If the peritoneum is inadvertently perforated, leading to the ingress of retroperitoneal gas into the peritoneal cavity, it can compromise the supportive configuration of the “peritoneal dome” and subsequently eliminate effective operating space. Drawing from our experiences with TEP surgery for hernia[28], we have found that even in cases of unintentional retroperitoneal injury, inserting a pneumoperitoneum needle into the abdominal cavity for gas evacuation can enable the successful continuation of retroperitoneal surgery.

In summary, this study investigates a novel surgical strategy for sigmoid colon tumors that shows promising early clinical efficacy and safety within a limited sample size. However, as a retrospective analysis, the quality of clinical evidence is limited, and the applicability of this novel “peritoneal dome” supported surgical strategy remains to be fully established. Therefore, further randomized controlled trials are necessary to assess its effectiveness and potential value in clinical settings.

## Data Availability

All data produced in the present study are available upon reasonable request to the authors.

